# ^99^mTc-FAPI SPECT/CT for Evaluation of Myocardial Fibrosis and Cardiac Function in Valvular Heart Disease

**DOI:** 10.1101/2025.04.17.25326016

**Authors:** Yu Zhang, Yue Tian, Guodong Guo, Haotian Zheng, Rongdong Xiao, Qi Xie, Yuanxiang Chen, Wenxin Chen, Chenshen Huang

## Abstract

**Background:** Active myocardial fibrosis plays a pivotal role in valvular heart disease (VHD) progression. While conventional imaging modalities primarily detect established fibrosis, the detection of active fibroblast activation remains challenging. ^99^mTc-FAPI SPECT/CT, offering advantages of wider availability and lower cost compared to PET imaging, represents a promising tool for evaluating active myocardial fibrosis.

**Methods:** We conducted a prospective study of 30 VHD patients who underwent ^99^mTc-FAPI SPECT/CT imaging with comprehensive clinical evaluation. Patients were categorized into FAPI-positive (n=24) and FAPI-negative (n=6) groups based on myocardial tracer uptake patterns. Thirteen FAPI-positive patients completed a three-month follow-up to assess therapeutic response.

**Results:** FAPI uptake parameters showed robust correlations with established myocardial injury markers (Hs-cTnI vs SUVmax: r=0.831, p<0.001; vs SUVmean: r=0.795, p<0.001) and disease severity indices (VHD staging: r=0.812, p<0.001). FAPI-positive patients demonstrated significantly higher atrial fibrillation prevalence (54.2% vs 0%, P=0.024). Post-treatment follow-up revealed significant improvements in FAPI parameters (SUVmax: 2.8±0.4 to 1.9±0.3, p<0.0001; SUVmean: 2.3±0.3 to 1.6±0.2, p<0.0001), accompanied by enhanced left ventricular ejection fraction (45.3±5.2% to 52.1±4.8%, p=0.0176) and NYHA functional status (p=0.0011), despite unchanged structural parameters.

**Conclusions:** This study demonstrates that ^99^mTc-FAPI SPECT/CT enables non-invasive visualization and quantification of active myocardial fibrosis in VHD patients. The strong associations between FAPI parameters and clinical indices, coupled with its ability to monitor therapeutic response, suggest its potential as a valuable tool for risk stratification and treatment optimization in VHD management.

**Trial registration:** ChiCTR2400094867. Registered 30 December 2024. Public site: https://www.chictr.org.cn/index.html.

## Introduction

Heart valve disease(VHD) is a prevalent condition that confers a substantial increase in mortality risk^1^. The underlying pathophysiology involves exposure of cardiac chambers to either pressure or volume overload, thereby initiating a cascade of maladaptive changes characterized by both reactive and replacement myocardial fibrosis, which ultimately leads to heart failure and arrhythmia^2,3^. Therefore, myocardial fibrosis, a hallmark of HVD progression, plays a pivotal role in the deterioration of cardiac function and serves as a critical determinant of clinical outcomes. Timely identification and precise evaluation of myocardial fibrosis are therefore imperative for improving patient outcomes. Currently, all widely utilized imaging modalities for assessing myocardial fibrosis have inherent limitations, as even the gold-standard technique of cardiac magnetic resonance with late gadolinium enhancement primarily measures extracellular expansion rather than direct fibrosis itself^3–5^. Moreover, the clinical implementation of MRI is significantly constrained by its prolonged acquisition time and contraindication in patients with cardiac pacemakers or other implantable devices, which substantially limits its widespread application in routine clinical practice.

Fibroblast Activation Protein (FAP), a cell surface serine protease predominantly expressed on activated fibroblasts, serves as both a specific molecular marker and a key mediator in the pathogenesis of tissue fibrosis. Recent study has demonstrated that FAP-targeted chimeric antigen receptor cells can reduce myocardial fibrosis and restore cardiac function after injury in a mouse model of heart failure ^6^. Concurrently, radiolabeled Fibroblast Activation Protein inhibitor (FAPI) tracers have emerged as a promising molecular imaging tool for the non-invasive assessment and quantification of myocardial fibrosis, enabling precise visualization of activated myocardial fibroblasts in various cardiovascular pathologies. Extensive clinical investigations have demonstrated the remarkable utility of 68Ga-FAPI PET/CT or PET/MR imaging in evaluating myocardial fibrosis across various conditions, including myocardial infarction^7,8^, hypertensive heart disease^9^, light-chain cardiac amyloidosis^10^, post-radiofrequency ablation status^11,12^, and idiopathic pulmonary arterial hypertension^13^.

However, PET/CT or PET/MR availability is limited in less developed countries, with fewer institutions possessing these devices compared to SPECT/CT. The restricted production of ^68^Ga and ^18^F, along with their short half-lives (^68^Ga: 67.71 min; ^18^F: 109.8 min), limits daily patient throughput. While SPECT systems have lower sensitivity and resolution than PET, recent SPECT/CT integration and absolute radioactivity quantification capabilities have significantly improved imaging quality^14–16^. ^99^mTc, available from ^99^Mo-^99^mTc generators, serves as an ideal SPECT radionuclide due to its longer half-life (361.2 min), cost-effectiveness, and widespread availability. While investigations utilizing ^99^mTc-FAPI SPECT/CT for myocardial fibrosis assessment remain limited, emerging evidence supports its clinical potential. Yu Liu and colleagues demonstrated that ^99^mTc-FAPI SPECT represents a promising imaging modality in idiopathic pulmonary fibrosis, with dosimetry studies confirming its favorable radiation profile compared to ^68^Ga- and ^18^F-labeled FAPI tracers, while maintaining an excellent safety profile^17^. Furthermore, Boqia Xie et al. revealed the remarkable sensitivity of ^99^mTc-HFAPi SPECT in detecting early-stage myocardial fibrotic processes in hypertensive conditions, even before the manifestation of conventional structural and functional cardiac alterations^18^.

However, the application of ^99^mTc-FAPI SPECT/CT for evaluating myocardial fibrosis in VHD remains unexplored, despite the practical advantages of ^99^mTc-based FAPI ligands in clinical settings. Therefore, we aimed to investigate the feasibility of ^99^mTc-FAPI SPECT/CT for the quantitative assessment of myocardial fibrosis in patients with VHD, potentially establishing a novel imaging paradigm for this specific cardiovascular condition.

## Materials and methods

### Study Population

Between January 2024 and October 2024, we retrospectively reviewed the medical records of patients who underwent ^99^mTc-FAPI SPECT/CT imaging at Fujian Provincial Hospital. Patients were considered eligible for inclusion if they met the following criteria: (1) age between 18 and 75 years, and (2) echocardiographic diagnosis of VHD according to current guidelines^19^. The exclusion criteria were: (1) documented history of myocardial infarction, (2) presence of malignant tumor, (3) pregnancy or lactation, and (4) women of childbearing potential. Clinical data were extracted from the hospital’s electronic medical record system using a standardized case report form by two independent investigators, and any disagreements were resolved through discussion with a third senior investigator. This retrospective study was approved by the Institutional Review Board of Fujian Provincial Hospital (approval number: K2024-12-075), with all procedures performed in accordance with the Declaration of Helsinki and institutional guidelines.

Baseline characteristics, including demographic data and cardiovascular risk factors, were collected for all patients. These parameters comprised body mass index (BMI), dyslipidemia, hypertension, diabetes, smoking status, atrial fibrillation, coronary heart disease (CHD), serum creatinine, high-sensitivity cardiac troponin I (hs-cTnI), and N-terminal pro-brain natriuretic peptide (NT-proBNP). Cardiac function was evaluated using the New York Heart Association (NYHA) functional classification class (Class I: no limitation of physical activity; Class II: slight limitation of physical activity; Class III: marked limitation of physical activity; Class IV: symptoms at rest)^20^. According to current guidelines, valvular heart disease (VHD) progression can be stratified into four distinct stages: Stage A represents individuals at risk for developing VHD but without detectable valvular alterations; Stage B encompasses patients with progressive valve disease showing mild to moderate severity without ventricular dysfunction; Stage C indicates severe asymptomatic valve disease, potentially accompanied by early ventricular dysfunction; and Stage D characterizes patients with severe symptomatic valve disease with advanced ventricular dysfunction, representing the end-stage of disease progression^19^. The New York Heart Association (NYHA) functional classification and valvular heart disease (VHD) staging were independently assessed by three experienced cardiologists (each with >5 years of clinical experience).

### 99mTc-Labeled SPECT/CT image acquisition

#### Radiopharmaceutical preparation

The FAPI precursor [HYNIC-(PEG4-OncoFAP)2] was supplied by the Medical Isotopes Research Center, Peking University(Beijing, China) for research and development. ^99^mTcO4− was procured from Guangdong CI Pharmaceutical Co., Fujian, China. For ^99^mTc radio labelling, 1 mL of 740-1369 MBq (20-37 mCi) of ^99^mTcO4− saline solution was added to 25 μg of HYNIC-(PEG4-OncoFAP)2 and then incubated at 100 for 20 minutes. The radiochemical purity of the final product exceeded 95%.

### SPECT/CT imaging

For ^99^mTc-FAPI SPECT/CT, patients received an intravenous injection of 925 MBq (25 mCi) of ^99^mTc-HOFAP2. One hour post-injection, regional (heart) SPECT/CT were conducted using the Discovery NM/CT 670Pro (GE, USA) equipped with low-energy, high-resolution collimators. The imaging protocol was as follows: for planar imaging, the peak energy was set at 140 keV (^99^mTc), with a scan velocity of 15 cm/min in a 256×1025 matrix. For regional SPECT/CT: the camera matrix size was 128×128; zoom factor, 1.0; rotation, 360°; 30 s/frame over 60 frames. A low-dose CT protocol (130 keV, 60 mA) was utilized.

### Imaging analysis

All images were independently analyzed by three experienced cardiac nuclear medicine physicians using a Xeleris Functional Imaging Workstation (Version 4.0; GE Healthcare). Differences in the determinations were discussed until a consensus was reached.(1) Visual analysis: FAPI uptake of myocardium was evaluated at visual inspection. FAPI uptake in the myocardium higher than in the adjacent blood pool was defined as abnormal. (2) Quantitative assessment: a volume of interest was manually drawn around the myocardium with FAPI uptake, and a region grow algorithm with a threshold of 40% of the maximum uptake was set to determine the FAPI-avid myocardium. Within this volume of interest, standard uptake value (SUV) was automatically derived, and the target-to-background ratio was calculated by dividing myocardial SUV by blood pool (superior vena cava) mean standardized uptake value to indicate the FAPI intensity. The raw data of FAPI images were further processed using QPS software (version 3.1; Cedars-Sinai Medical Center) and polar plot was generated. The FAPI extent (the percent of FAPI-avid myocardium of the LV) was obtained from the polar plot using the defined threshold. Maximum standardized uptake value (SUVmax) representing peak myocardial fibroblast activation intensity, mean standardized uptake value (SUVmean) reflecting overall fibrosis degree, and Extent indicating the percentage of myocardium with active fibrosis.

### Echocardiographic

Transthoracic echocardiography was performed using EPIQ CVx cardiology ultrasound system (Philips Medical Systems, Andover, Massachusetts, USA) to assess cardiac structure and function. The following parameters were measured: interventricular septum (IVS) thickness, left ventricular diastolic diameter (LVDD), Left atrium(LA) diameter, left ventricular posterior wall (LVPW) thickness, left ventricular end-diastolic volume (LVEDV), and left ventricular ejection fraction (LVEF).

### Treatment and Follow-up

Treatment strategies were individualized according to established clinical guidelines, encompassing both surgical interventions or medical therapy based on comprehensive clinical assessment and patient preferences. Follow-up evaluations were systematically conducted at 3 months post-treatment, with echocardiography for cardiac function assessment and ^99^mTc-FAPI SPECT/CT for myocardial fibrosis evaluation.

### Statistical Analysis

Statistical analyses were performed using R software (version 4.2.0; R Foundation for Statistical Computing, Vienna, Austria). The Shapiro-Wilk test was employed to assess data normality. Continuous variables were presented as mean ± standard deviation or median (interquartile range) for normal and non-normal distributions, respectively. Between-group comparisons were conducted using independent t-tests or Mann-Whitney U tests as appropriate. Categorical variables were expressed as numbers (percentages) and compared using Fisher’s exact test given the small sample size.

To comprehensively evaluate the relationships between FAPI parameters and clinical indicators, both Pearson and Spearman correlation analyses were performed. Longitudinal changes were assessed using paired t-tests or Wilcoxon signed-rank tests for continuous variables, and McNemar test for categorical variables. Two-sided P values <0.05 were considered statistically significant.

## Results

### Patient Characteristics and Baseline Demographics

During the study period, a total of 446 patients underwent 99mTc-FAPI SPECT/CT imaging, of whom 33 initially met the inclusion criteria. Three patients were subsequently excluded: two due to documented history of myocardial infarction and one due to the presence of malignant tumor. Ultimately, 30 patients were included in the final analysis (Figure 1). The study population consisted of 16 males (53.3%) and 14 females (46.7%), with a mean age of 59.3 ± 12.15 years. The baseline characteristics of the study population are summarized in Table 1. Patients were stratified into positive (n=24) and negative (n=6) groups, showing comparable baseline characteristics including age (59.625 ± 12.97 vs 58 ± 8.90 years, P = 0.775), BMI (22.84 ± 4.89 vs 22.154 ± 1.29 kg/m², P = 0.739), and traditional cardiovascular risk factors such as hypertension (45.8% vs 50%, P = 1.000), dyslipidemia (20.8% vs 50%, P = 0.300), and diabetes mellitus (8.3% vs 16.7%, P = 0.501). Notably, atrial fibrillation demonstrated significantly higher prevalence in the positive group (54.2% vs 0%, P = 0.024).

**Table 1.**
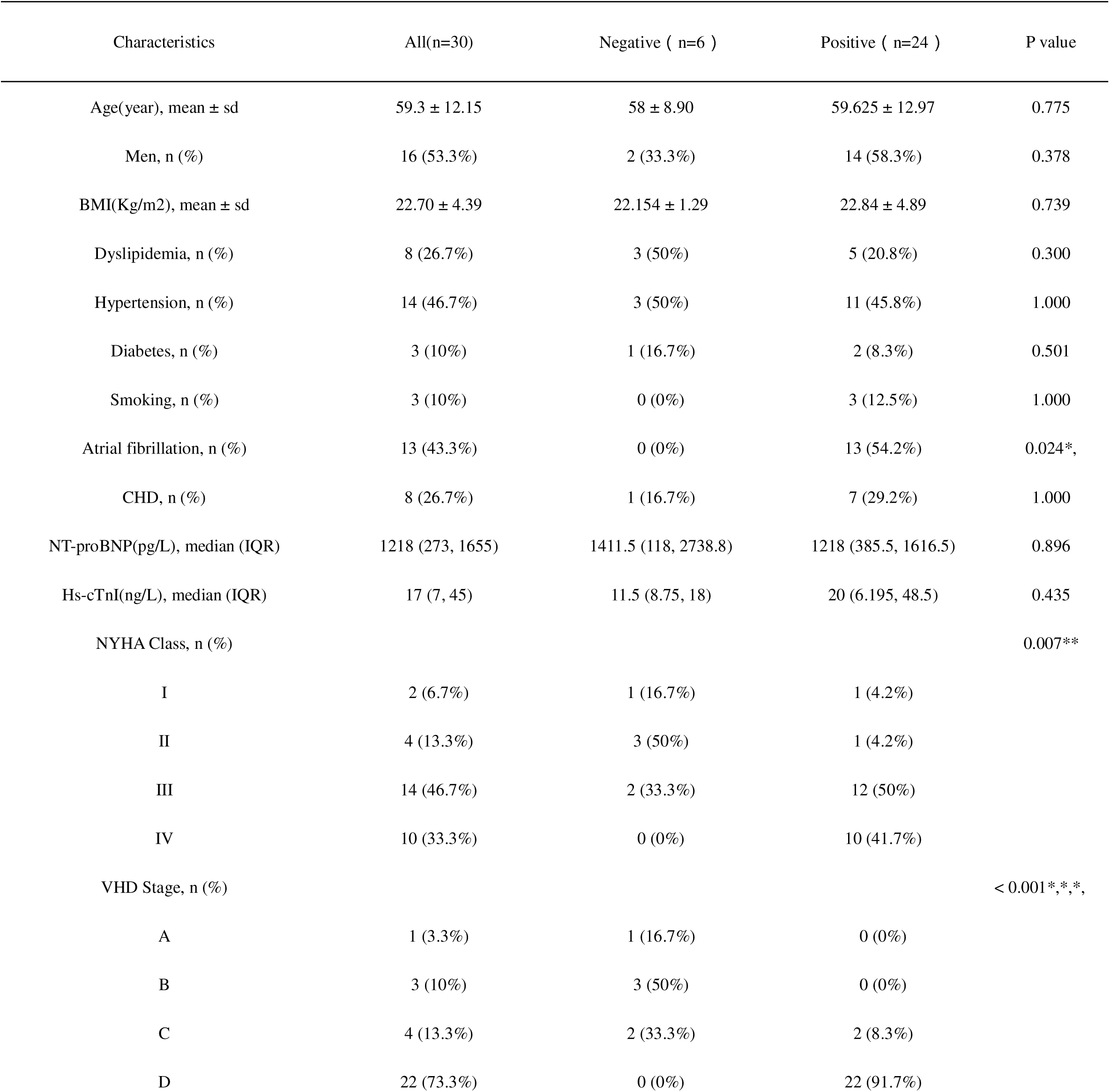
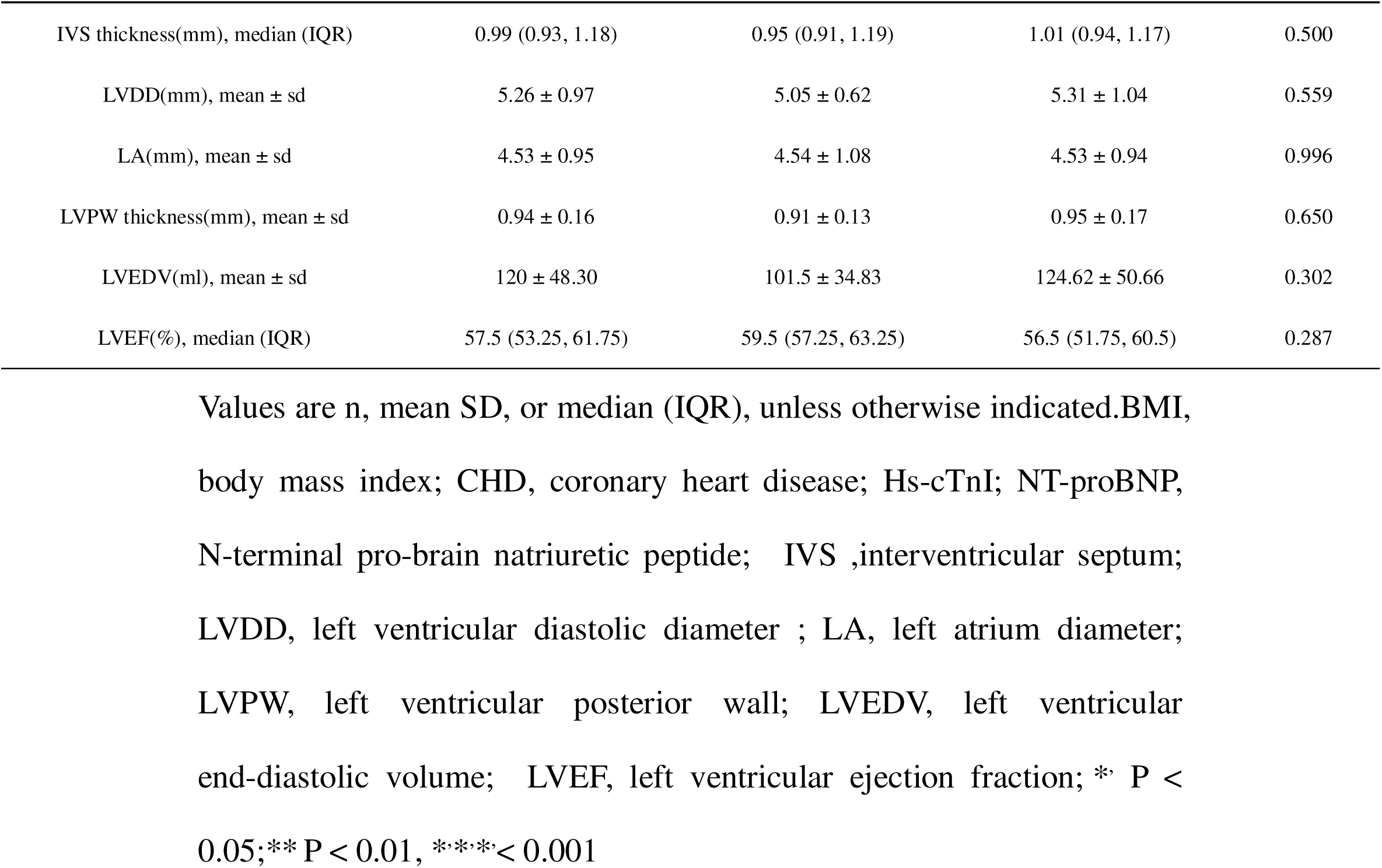
Baseline Demographic and Clinical Characteristics of Patients With Heart Valve Disease (n=30)

| Characteristics | All(n=30) | Negative ( n=6 ) | Positive ( n=24 ) | P value |
| --- | --- | --- | --- | --- |
| Age(year), mean ± sd | 59.3 ± 12.15 | 58 ± 8.90 | 59.625 ± 12.97 | 0.775 |
| Men, n (%) | 16 (53.3%) | 2 (33.3%) | 14 (58.3%) | 0.378 |
| BMI(Kg/m2), mean ± sd | 22.70 ± 4.39 | 22.154 ± 1.29 | 22.84 ± 4.89 | 0.739 |
| Dyslipidemia, n (%) | 8 (26.7%) | 3 (50%) | 5 (20.8%) | 0.300 |
| Hypertension, n (%) | 14 (46.7%) | 3 (50%) | 11 (45.8%) | 1.000 |
| Diabetes, n (%) | 3 (10%) | 1 (16.7%) | 2 (8.3%) | 0.501 |
| Smoking, n (%) | 3 (10%) | 0 (0%) | 3 (12.5%) | 1.000 |
| Atrial fibrillation, n (%) | 13 (43.3%) | 0 (0%) | 13 (54.2%) | 0.024*, |
| CHD, n (%) | 8 (26.7%) | 1 (16.7%) | 7 (29.2%) | 1.000 |
| NT-proBNP(pg/L), median (IQR) | 1218 (273, 1655) | 1411.5 (118, 2738.8) | 1218 (385.5, 1616.5) | 0.896 |
| Hs-cTnI(ng/L), median (IQR) | 17 (7, 45) | 11.5 (8.75, 18) | 20 (6.195, 48.5) | 0.435 |
| NYHA Class, n (%) |  |  |  | 0.007** |
| I | 2 (6.7%) | 1 (16.7%) | 1 (4.2%) |  |
| II | 4 (13.3%) | 3 (50%) | 1 (4.2%) |  |
| III | 14 (46.7%) | 2 (33.3%) | 12 (50%) |  |
| IV | 10 (33.3%) | 0 (0%) | 10 (41.7%) |  |
| VHD Stage, n (%) |  |  |  | < 0.001*,*,*, |
| A | 1 (3.3%) | 1 (16.7%) | 0 (0%) |  |
| B | 3 (10%) | 3 (50%) | 0 (0%) |  |
| C | 4 (13.3%) | 2 (33.3%) | 2 (8.3%) |  |
| D | 22 (73.3%) | 0 (0%) | 22 (91.7%) |  |
| IVS thickness(mm), median (IQR) | 0.99 (0.93, 1.18) | 0.95 (0.91, 1.19) | 1.01 (0.94, 1.17) | 0.500 |
| LVDD(mm), mean ± sd | 5.26 ± 0.97 | 5.05 ± 0.62 | 5.31 ± 1.04 | 0.559 |
| LA(mm), mean ± sd | 4.53 ± 0.95 | 4.54 ± 1.08 | 4.53 ± 0.94 | 0.996 |
| LVPW thickness(mm), mean ± sd | 0.94 ± 0.16 | 0.91 ± 0.13 | 0.95 ± 0.17 | 0.650 |
| LVEDV(ml), mean ± sd | 120 ± 48.30 | 101.5 ± 34.83 | 124.62 ± 50.66 | 0.302 |
| LVEF(%), median (IQR) | 57.5 (53.25, 61.75) | 59.5 (57.25, 63.25) | 56.5 (51.75, 60.5) | 0.287 |

Values are n, mean SD, or median (IQR), unless otherwise indicated.BMI, body mass index; CHD, coronary heart disease; Hs-cTnI; NT-proBNP, N-terminal pro-brain natriuretic peptide; IVS ,interventricular septum; LVDD, left ventricular diastolic diameter ; LA, left atrium diameter; LVPW, left ventricular posterior wall; LVEDV, left ventricular end-diastolic volume; LVEF, left ventricular ejection fraction; \* P < 0.05;\*\* P < 0.01, \*\*\*P < 0.001

### Clinical Parameters and Disease Severity

Table 1 shows disease severity assessment revealed significant differences between groups. The distribution of NYHA functional classification showed marked variation (P = 0.007), with the positive group demonstrating more advanced symptoms (Class III: 50%, Class IV: 41.7%) compared to the negative group where most patients were Class I (16.7%) or Class II (50%). VHD staging showed significant differences between groups (P < 0.001). In the positive group, 91.7% of cases were classified as Stage D and 8.3% as Stage C, while the negative group showed a different distribution with 16.7% in Stage A, 50% in Stage B, and 33.3% in Stage C. Cardiac biomarkers, including NT-proBNP [1218 (385.5, 1616.5) vs 1411.5 (118, 2738.8) pg/L, P = 0.896] and hs-cTnI [20 (6.195, 48.5) vs 11.5 (8.75, 18) ng/L, P = 0.435], showed no significant differences between groups. Echocardiographic parameters, including LVEF [56.5 (51.75, 60.5)% vs 59.5 (57.25, 63.25)%, P = 0.287] and cardiac dimensions, were comparable between groups. Echocardiographic parameters were comparable between groups, with no significant differences in LVEF [positive: 56.5 (51.75, 60.5)% vs negative: 59.5 (57.25, 63.25)%, P = 0.287], cardiac dimensions including LVDD (5.31 ± 1.04 vs 5.05 ± 0.62 mm, P = 0.559), LA diameter (4.53 ± 0.94 vs 4.54 ± 1.08 mm, P = 0.996), and other structural measurements.

### FAPI Parameters with Clinical Indicators

We performed comprehensive correlation analyses to evaluate the relationships between imaging parameters and clinical indices (Figure 2). Pearson correlation analysis (Figure 2A) revealed strong positive correlations between cardiac troponin I (hs-cTnI) and both SUVmax (r = 0.831, P < 0.001) and SUVmean (r = 0.795, P < 0.001). The interventricular septum (IVS) thickness also demonstrated significant positive correlations with SUVmax (r = 0.698, P < 0.001) and SUVmean (r = 0.658, P < 0.001). Other cardiac structural and functional parameters (LVDD, LA, LVPW, LVEDV, LVEF, and NT-proBNP) showed no significant correlations with imaging parameters (all P > 0.05).

**Figure 1.**
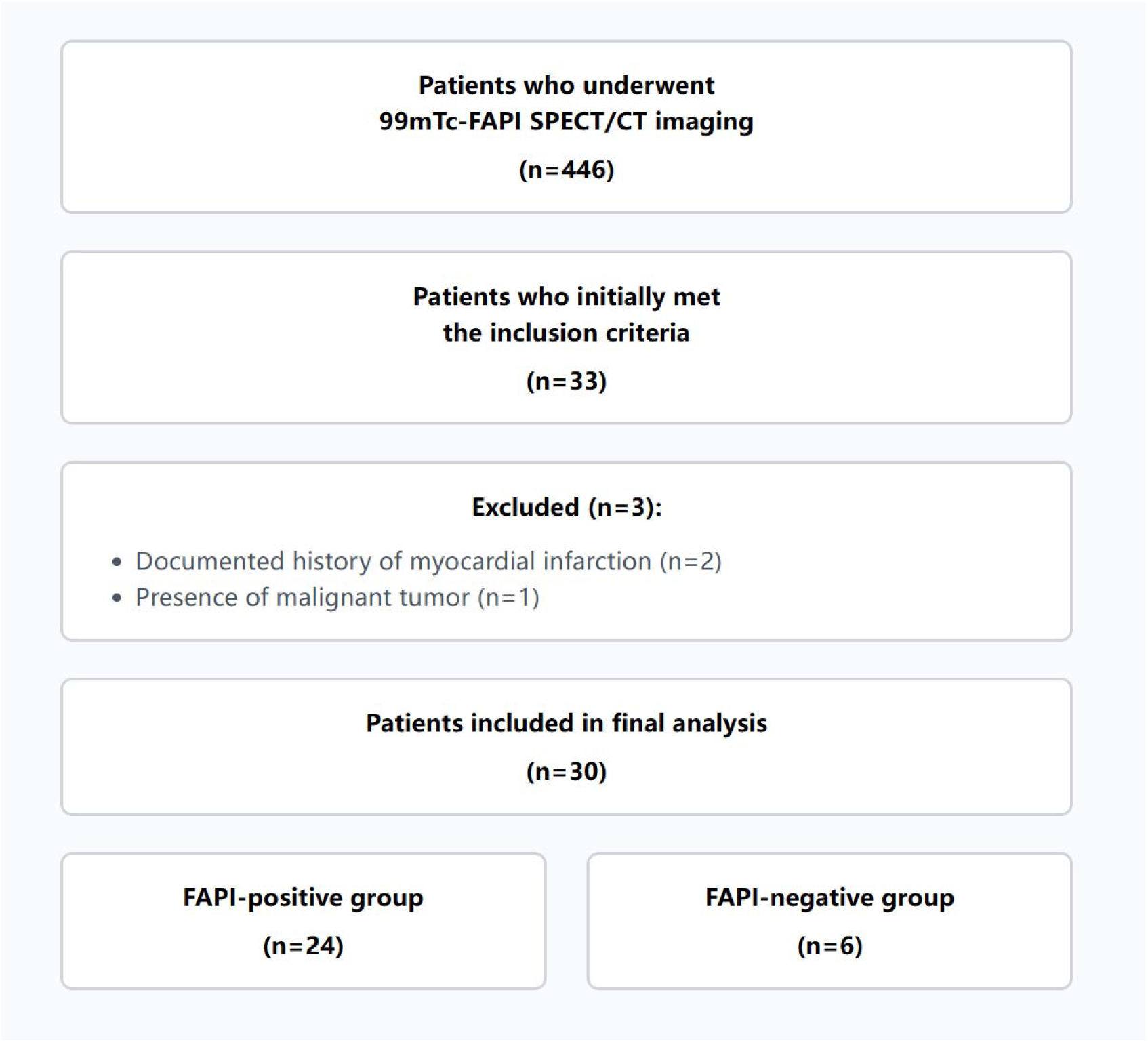
Flow diagram of patient recruitment.

**Figure 2.**
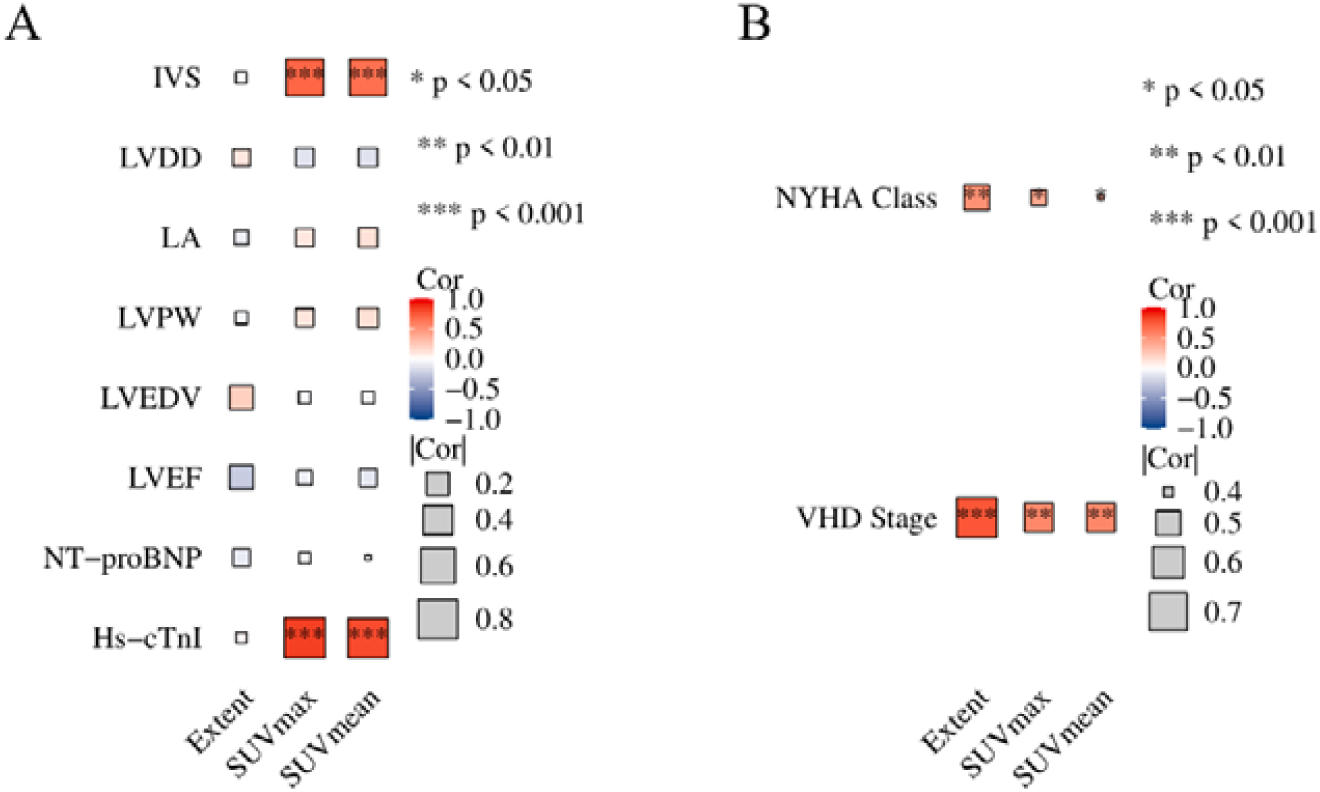
Correlation analysis between imaging parameters and clinical indices.(A) Correlation analyses between FAPI imaging parameters and clinical indices. Pearson correlation analysis revealed strong positive correlations between Hs-cTnI and both SUVmax (r = 0.831, P < 0.001) and SUVmean (r = 0.795, P < 0.001), while IVS thickness showed significant correlations with SUVmax (r = 0.698, P < 0.001) and SUVmean (r = 0.658, P < 0.001). (B)Spearman analysis demonstrated strong correlations between the Extent parameter and both NYHA classification (ρ = 0.570, P = 0.001) and VHD staging (ρ = 0.849, P < 0.001), with SUVmax and SUVmean showing moderate correlations with VHD staging (ρ = 0.424-0.445, P < 0.05). Color scale represents correlation coefficient (P) values. (*P<0.05, **P<0.01, ***P<0.001).

Spearman correlation analysis (Figure 2B) demonstrated significant associations between disease severity indices and imaging parameters. The Extent parameter showed strong correlations with both NYHA functional classification (ρ = 0.570, P = 0.001) and VHD staging (ρ = 0.849, P < 0.001). Additionally, SUVmax and SUVmean exhibited moderate correlations with VHD staging (ρ = 0.424, P = 0.020 and ρ = 0.445, P = 0.014, respectively).

### Treatment Response and Follow-up Outcomes

During the 3-month follow-up period, 18 patients (78.3%) completed follow-up evaluation, including 5 cases (27.8%) in the FAPI-negative group and 13 cases (72.2%) in the positive group. The negative group maintained negative FAPI uptake at follow-up. In the positive group, significant improvements were observed across all FAPI imaging parameters. As illustrated in Figure 3A-C, the extent of FAPI uptake decreased significantly (P=0.033), accompanied by marked reductions in both SUVmax (P<0.0001) and SUVmean (P<0.0001). Echocardiographic assessment (Figure 3D-H) showed no significant changes in cardiac structural parameters, including IVS, LVDD, LA, LVPW, and LVEDV over the follow-up period. However, LVEF demonstrated significant improvement (P=0.0176; Figure 3I). Notably, NYHA functional classification showed marked improvement (P=0.0011; Figure 3J).

**Figure 3.**
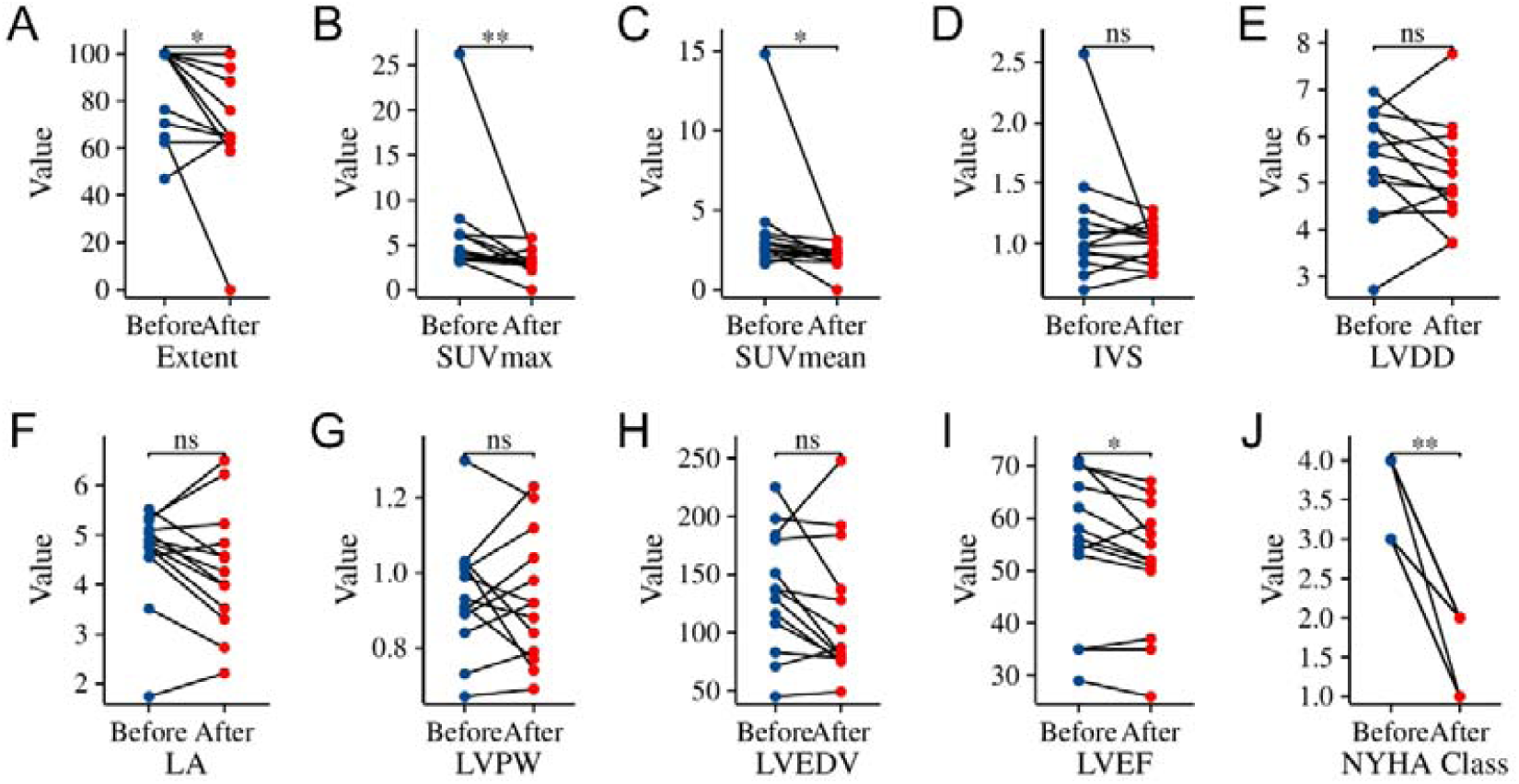
Changes in FAPI imaging parameters, echocardiographic indices, and NYHA classification during 3-month follow-up in FAPI-positive VHD patients (n=13). (A-C) FAPI-related parameters showing significant improvements in Extent (P=0.033), SUVmax (P<0.0001), and SUVmean (P<0.0001). (D-I) Echocardiographic parameters including IVS, LVDD, LA, LVPW, and LVEDV showing no significant changes, while LVEF demonstrating significant improvement (P=0.0176). (J) Significant improvement in NYHA classification (P=0.0011). Box plots show median, interquartile range, and range; dots represent individual values. *P<0.05, **P<0.01, ***P<0.001.

Figure 4 demonstrates representative pre- and post-operative ^99^mTc-FAPI SPECT/CT images from a 57-year-old female patient with severe rheumatic mitral stenosis who underwent mechanical mitral valve replacement. The 3-month follow-up images showed complete resolution of myocardial FAPI uptake following surgery, providing visual evidence of the therapeutic response.

**Figure 4.**
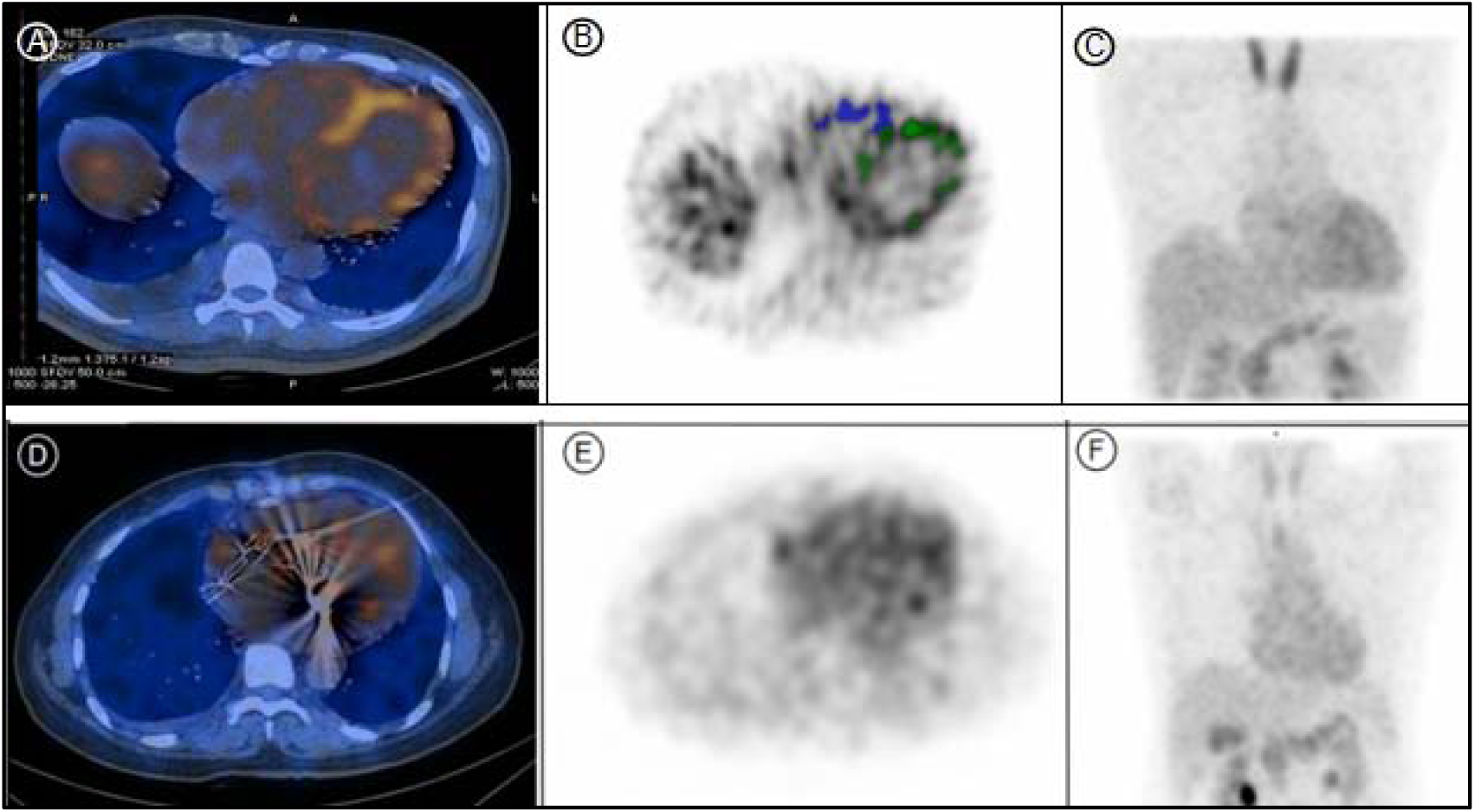
Representative ^99^mTc-FAPI SPECT/CT images of a 57-year-old female patient with severe rheumatic mitral stenosis who underwent mechanical mitral valve replacement. Pre-operative images (A-C) and post-operative images (D-F) demonstrate complete resolution of myocardial FAPI uptake following surgery. (A, D) Axial fused SPECT/CT images showing anatomical localization of tracer uptake. (B, E) Corresponding SPECT-only axial images with color-coded regions of interest, where green annotations indicate left ventricular myocardial uptake and blue annotations highlight right ventricular myocardial uptake in pre-operative images, with no detectable uptake post-operatively. (C, F) Maximum intensity projection (MIP) images providing three-dimensional visualization of tracer distribution, confirming the absence of pathological FAPI uptake after valve replacement.

## Discussion

In this study, we demonstrated for the first time that ^99^mTc-FAPI SPECT/CT imaging provides valuable information for assessing myocardial fibrosis in VHD patients, offering several novel insights into disease mechanisms and potential clinical applications.

Myocardial fibrosis, characterized by excessive extracellular matrix deposition and cardiac remodeling, represents a fundamental pathophysiological mechanism in cardiovascular disease progression^21,22^. This pathological process has been implicated in the progression of VHD and subsequent myocardial injury, potentially contributing to cardiac dysfunction and atrial fibrillation ^2,23–26^. In our preliminary study, FAPI-PET imaging analysis suggested possible mechanistic relationships. Initial quantitative analysis revealed correlations between myocardial injury marker (Hs-cTnI) and FAPI uptake parameters (SUVmax: r = 0.831, p < 0.001; SUVmean: r = 0.795, p < 0.001). FAPI parameters also showed associations with clinical severity indicators, including NYHA functional class (r = 0.756, p < 0.001) and VHD staging (r = 0.812, p < 0.001). In our small cohort, we observed a higher prevalence of AF in FAPI-positive patients compared to FAPI-negative counterparts (54.2% vs 0%, P = 0.024). These preliminary findings suggest that FAPI-PET imaging may have potential as a non-invasive tool for evaluating myocardial fibrosis in VHD patients.

Our preliminary findings suggest that ^99^mTc-FAPI SPECT/CT may provide complementary information about active fibrosis that differs from the structural changes detected by conventional cardiac MRI. This observation aligns with recent evidence suggesting that molecular imaging techniques may identify early pathological changes before structural alterations become apparent^27,28^. In FAPI-positive VHD patients (n=13), three-month follow-up demonstrated promising therapeutic responses: FAPI imaging parameters showed significant reductions in Extent (P=0.033), SUVmax (P<0.001), and SUVmean(P<0.001). While conventional structural echocardiographic parameters (IVS, LVDD, LA, LVPW, and LVEDV) remained stable, left ventricular ejection fraction improved significantly (P=0.0176), accompanied by marked improvement in NYHA functional classification (P=0.0011). These findings suggest that FAPI imaging may serve as a sensitive tool for early detection of therapeutic response and cardiac functional recovery, potentially facilitating timely intervention strategies.

However, several limitations should be acknowledged. First, our sample size was relatively small, necessitating larger multicenter studies for result validation. Second, the follow-up period was limited, preventing assessment of long-term prognostic value. Third, SPECT’s inherent spatial resolution limitations may affect detection of small lesions. Fourth, direct comparison with other molecular imaging techniques was not performed.

In conclusion, our study establishes ^99^mTc-FAPI SPECT/CT as a promising tool for assessing myocardial fibrosis in VHD patients, potentially offering earlier detection and better monitoring of disease progression than conventional imaging. While further validation is needed, this technique may represent a significant advance in the molecular imaging of cardiac fibrosis, with important implications for patient care. As technology evolves and clinical experience accumulates, FAPI imaging has the potential to become an integral component in the management of VHD patients, particularly in early intervention and therapeutic monitoring strategies.

## Data Availability

All data produced in the present study are available upon reasonable request to the authors

## Statements & Declarations

### Author Contributions

Conceptualization, Y.C., and C.H; Methodology, Y.Z., Y.T., G.G.; Literature Research, Y.Z., Y.T., H.Z., R.X., C.H and W.C.; Data Acquisition, Y.Z., Y.T., H.Z., and Y.C.; Data Analysis & Interpretation, Y.C.; Writing—Original Draft, Y.Z., C.H., and Y.C.; Writing—Review & Editing, R.X., Q.X and W.C.; All authors read and approved the submitted version of manuscript.

### Data Availability

This paper does not report the original code. Any additional information required to reanalyze the data reported in this paper is available from the lead contact upon request.

### Ethics approval

This retrospective study was approved by the Institutional Review Board of Fujian Provincial Hospital (approval number: K2024-12-075). All procedures were performed in accordance with the Declaration of Helsinki and institutional guidelines.

### Consent to participate

For all research involving human subjects, freely-given, informed consent to participate in the study must be obtained from participants (or their parent or legal guardian in the case of children under 16) and a statement to this effect should appear in the manuscript.

### Example statement

Due to the retrospective nature of this study, the requirement for individual informed consent was waived by the Institutional Review Board of Fujian Provincial Hospital.

### Consent to publish

All authors consent to the publication of the manuscript and verify that the data presented in this study adheres to journal policies. Patient identifiable information has been anonymized in accordance with ethical standards.

#### Acknowledgments

The authors thank Qi Huang (Shanghai Tongji University, Shanghai, China) and Hong Chen (Fujian Provincial Hospital, Fuzhou, China) for generously sharing their experience.

## Additional resources

Trial registration: ChiCTR2400094867. Registered 30 December 2024. Public site: https://www.chictr.org.cn/index.html.

